# Dynamics of spinal fluid immune cell alterations following cladribine tablet treatment in multiple sclerosis

**DOI:** 10.1101/2025.02.22.25322629

**Authors:** Roman A. Smirnov, Claudia Cantoni, Kenneth Lee, Rea A. Agnihotri, Robert C. Axtell, Amber Salter, Samantha Lancia, Michael A. Paley, Brooke Hayward, Julie Korich, Emily Evans, Gabriel Pardo, Olaf Stuve, Amit Bar-Or, Maxim N. Artyomov, Anne H. Cross, Brian T. Edelson, Gregory F. Wu

**Affiliations:** Department of Pathology and Immunology, Washington University in St. Louis, St. Louis, MO 63110, USA; Barrow Neurological Institute; 2910 N. 3rd Ave., Phoenix, AZ 85013 USA; Department of Neurology, Washington University in St. Louis, St. Louis, MO 63110, USA; Department of Arthritis and Clinical Immunology Research, Oklahoma Medical Research Foundation, Oklahoma City, OK, USA; Multiple Sclerosis Center of Excellence, Oklahoma Medical Research Foundation, Oklahoma City, OK 73104, USA; Department of Neurology, University of Texas Southwestern Medical Center, Dallas, TX 75390, USA; Department of Medicine, Washington University in St. Louis, St. Louis, MO 63110, USA; EMD Serono, Boston, MA 02210, USA; The Center for Neuroinflammation and Experimental Therapeutics and the Department of Neurology, University of Pennsylvania, Philadelphia, PA, 19104, USA

## Abstract

**Background and Objectives:** Oral cladribine tablet (CladT) therapy is efficacious for relapsing multiple sclerosis (MS). However, the mechanisms by which cladribine exerts benefit in MS remain unclear, particularly regarding its effects on compartmentalized inflammation within the cerebrospinal fluid (CSF).

**Methods:** Transcriptional profiles along with T and B lymphocyte receptor repertoires from CSF and blood were obtained by single cell sequencing methods from a single site participating in a phase IV clinical trial investigating the impact of cladribine treatment for MS. All subjects provided blood and CSF samples immediately before starting CladT therapy and were randomized to also provide samples at either five weeks, ten weeks, one year, or two years post-CladT therapy. Thirty-four samples from 13 individuals with relapsing MS before and after treatment were available to test the hypothesis that CladT alters the composition and phenotype of lymphocytes in the CSF.

**Results:** We found that treatment with CladT profoundly altered cellular composition, but not the transcriptional phenotype, of immune cells in the CSF. In particular, we identified a reduction of switched memory B cells but recovery of naive B cells in the CSF, similar to our findings in blood. Additionally, populations of CD4 Treg cells emerged early after CladT therapy and remained elevated one year later in the CSF, but not in the blood. Antigen receptor sequencing revealed a moderate decrease in numbers of large clonally expanded CD8 T cell clones (>10 cells/clone) primarily in the CSF, but also in the blood post-CladT treatment.

**Discussion:** Our results identified unique cellular dynamics and changes in T and B cell clonality in both tissues which can potentially explain long-term beneficial effects of CladT therapy in MS, including preservation of immune function and relatively low number of side effects. Altogether, this study demonstrates that CladT treatment had a substantial impact not only on blood, but also on the CSF compartment highlighting the importance of cross-tissue analysis for better understanding of effect and the mechanism of action of DMTs.

## Introduction

Despite numerous disease-modifying therapies (DMTs) available for treating relapsing multiple sclerosis (MS), the exact pathogenesis of the disease is still unknown^1,2^. The varying success of different DMTs with distinctive mechanisms of action has led to the identification of several immunologic pathways involved in MS, including lymphocyte trafficking^3,4^, B cell antigen presentation^5,6^, and interferon expression^7,8^. Cladribine disrupts DNA metabolism causing immune cell death, and oral cladribine tablet (CladT) administration is efficacious in reducing disease activity in relapsing MS^9–11^. Treatment with CladT induces a transient leukopenia that is proposed to reduce neuroinflammation by removal of deleterious lymphocytes and subsequent repopulation of the lymphocyte compartment in the periphery with tolerogenic immune cells^12–14^. How CladT treatment, including its effects on peripheral immune cell populations, affects immune cell subsets within the central nervous system (CNS) is currently unknown.

An important aspect of MS involves compartmentalized inflammation, with CSF biomarkers offer a window into mechanisms underlying CNS injury and therapeutic potential for various stages of disease^15^. While several soluble inflammatory markers associated with MS disease activity and response to therapy have been pursued^16,17^, cellular immune CSF biomarkers of MS treatment are just starting to emerge^18,19^. Traditionally, access to, and viability of, the limited number of cells have stymied efforts to utilize the cellular component of CSF as a reliable biomarker for DMT effects in MS.

Advancements in the characterization of immune cells in the steady-state as well as in autoimmune and neurologic diseases have been made using single-cell RNA-sequencing (scRNA-seq)^20–22^. The power of scRNA-seq lies in its high-throughput, high-resolution nature of gene expression profiling. Several recent studies have demonstrated the diversity of immune cells contained in the CSF of subjects with neurologic disease, including MS^23–27^. These have provided insight into the presence of various immune cell subsets unique to disease states. Transcriptomic profiling of CSF offers an additional opportunity to characterize CNS compartmentalized immune cell features before and after DMT administration for MS which could possibly identify immunologic changes associated with benefit in addition to uncovering potential insights into DMT mechanisms of action.

We therefore profiled samples from CSF and peripheral blood mononuclear cells (PBMCs) from at pre-specified time points after CladT treatment. All samples were analyzed using scRNA-seq inclusive of single cell T cell receptor (scTCR) and single cell B cell receptor (scBCR) sequencing. A total of 34 samples from 13 patients with relapsing MS yielded a dataset spanning 72,547 cells with 32 biologically distinct cell types. Samples from CSF and blood allowed us to compare CladT-mediated cell dynamics. CSF B cell dynamics were characterized by long-term depletion of switched memory B cell subsets and rapid recovery of naive B cells. Additionally, we identified a long-lasting, CSF-specific increase in CD4 Treg subsets after CladT therapy. We also found that CladT substantially reduced the number of large CD8 T cell clones (>10 cells/clone) predominantly in the CSF, but also in blood, which may contribute to amelioration of MS disease activity. Overall, our study provides comprehensive characterization of CladT-associated cellular remodeling in blood and CSF along with its effect on T and B cell clonality. These findings shed light on some possible immune mechanisms associated with the beneficial effects of CladT treatment in MS.

## Results

### Study design for transcriptional profiling of blood and CSF immune cells in CladT-treated MS patients

We used an unbiased approach to identify compartment-specific changes in the composition and clonality of immune cell types in the blood and CSF from MS patients associated with CladT therapy (**Figure 1A**). Accordingly, relapsing MS patients were enrolled in the collaborative phase IV clinical trial, Cladribine Tablets: Collaborative Study to Evaluate Impact on Central Nervous System Biomarkers in Multiple Sclerosis (CLOCK-MS). Participants provided CSF and blood samples prior to initiating CladT treatment and were then randomized to different times following treatment for obtaining second CSF sample. Six paired blood and CSF samples were collected from individual subjects before oral CladT treatment. Of these six individuals, four provided paired blood and CSF samples 5- or 10-weeks post-treatment or one year post-treatment (prior to re-dosing). An additional individual provided paired blood and CSF samples two years following CladT treatment initiation. Additional non-paired blood samples were obtained from nine subjects (**Table S1**). In total, 34 samples from 13 participants were collected over two years and analyzed by scRNA/scTCR/scBCR-seq using the 10x Genomics platform (**Figure 1A**). After initial quality control and individual sample preprocessing, 72,547 transcriptomes (49,249 PBMC and 23,298 CSF cells) were integrated into a common object (**Figure 1B**, **Table S1**). To define individual immune cell subpopulations, we utilized a hierarchical clustering approach on the merged 72,547 blood/CSF cell dataset. The major immune subsets of B, myeloid, natural killer (NK), CD4 and CD8 T cells were identified. These major populations were further subclustered into 32 distinct immune subpopulation from blood and CSF matching markers or transcriptional signatures of biologically well-defined subsets^28^ (**Figure 1B–E**). We then focused on each blood and CSF subcluster to explore CladT-mediated changes in cell type proportions as well as clonality of T and B cells.

**Figure 1.**
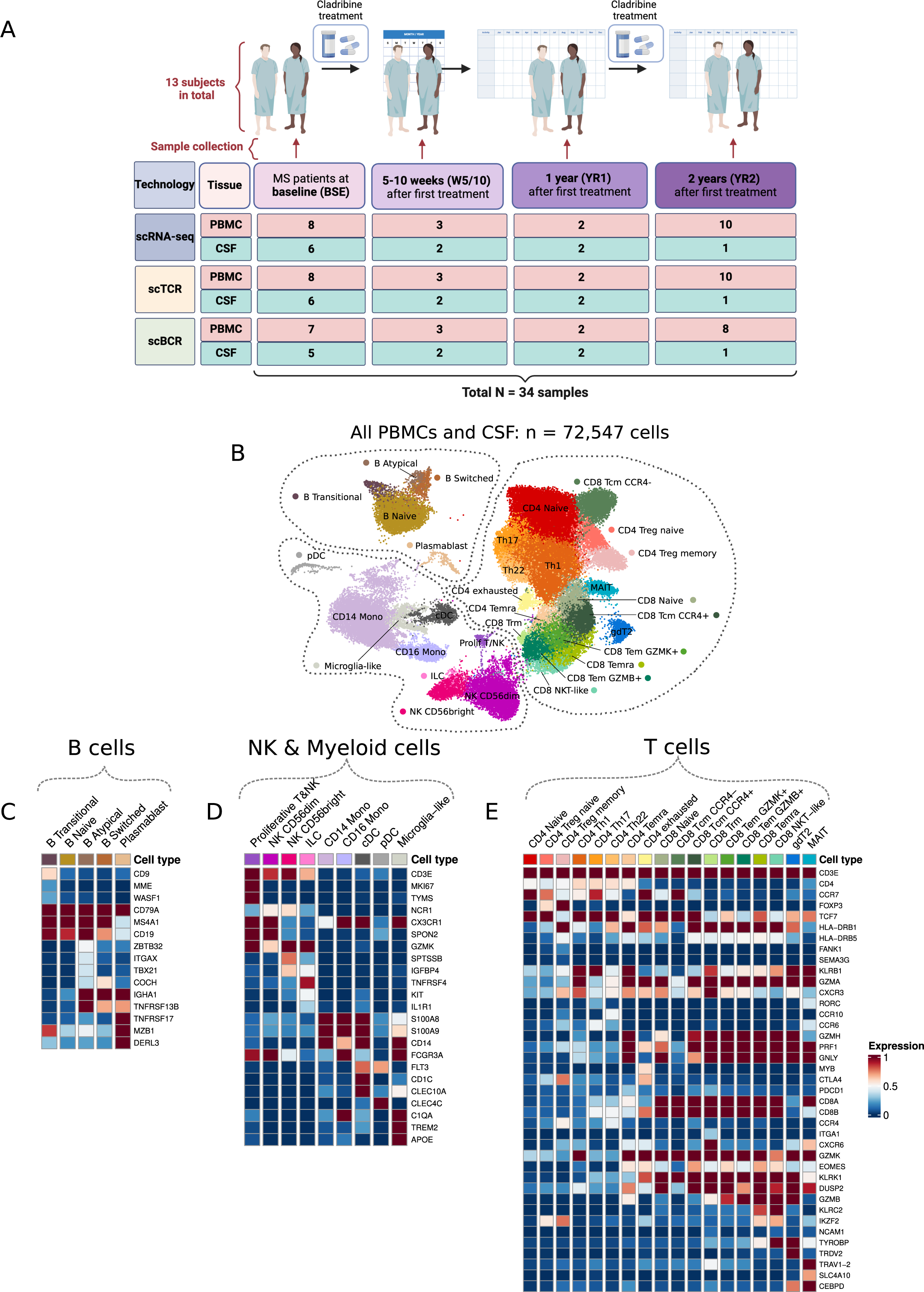
Defining immune cell subsets from PBMCs and CSF of MS subjects treated with CladT. (**A**) Schematic of the study design and sample cohort (image was created using Biorender).(B) UMAP of 72,547 cells collected from both PBMCs and CSF of 13 subjects. (**C–E**) Matrix plot of marker genes designating (**C**) B cell, (**D**) NK and Myeloid cell, and (**E**) T cell subsets.

### Remodeling of immune cell type composition in blood after CladT treatment

As a result of unbiased scRNA-seq analysis of 23 blood samples collected at different time points after CladT therapy (**Figure 1A**), the PBMC dataset was composed of 49,249 cells revealing 31 biologically distinct clusters (**Figure 2A**). As expected for blood, naive, switched memory and atypical memory B cells were drastically reduced in proportion early on after CladT treatment (**Figure 2B, C**). After 1 and 2 years of therapy, switched and atypical memory B cells were persistently depleted in blood, although this did not reach statistical significance at year 1 (**Figure 2D, E**). Conversely, naive B cells showed a significant increase in proportion at year 1 but were not significantly different from baseline two years after CladT therapy. Treatment with CladT reduced plasmablast numbers in blood after one year of therapy, but this difference did not reach statistical significance. The rapid response by B cells in the blood to CladT treatment are consistent with a recent report by Teschner et al^29^ and in line with the presumed mechanism of action on peripheral cell profiles after CladT therapy in MS.

**Figure 2.**
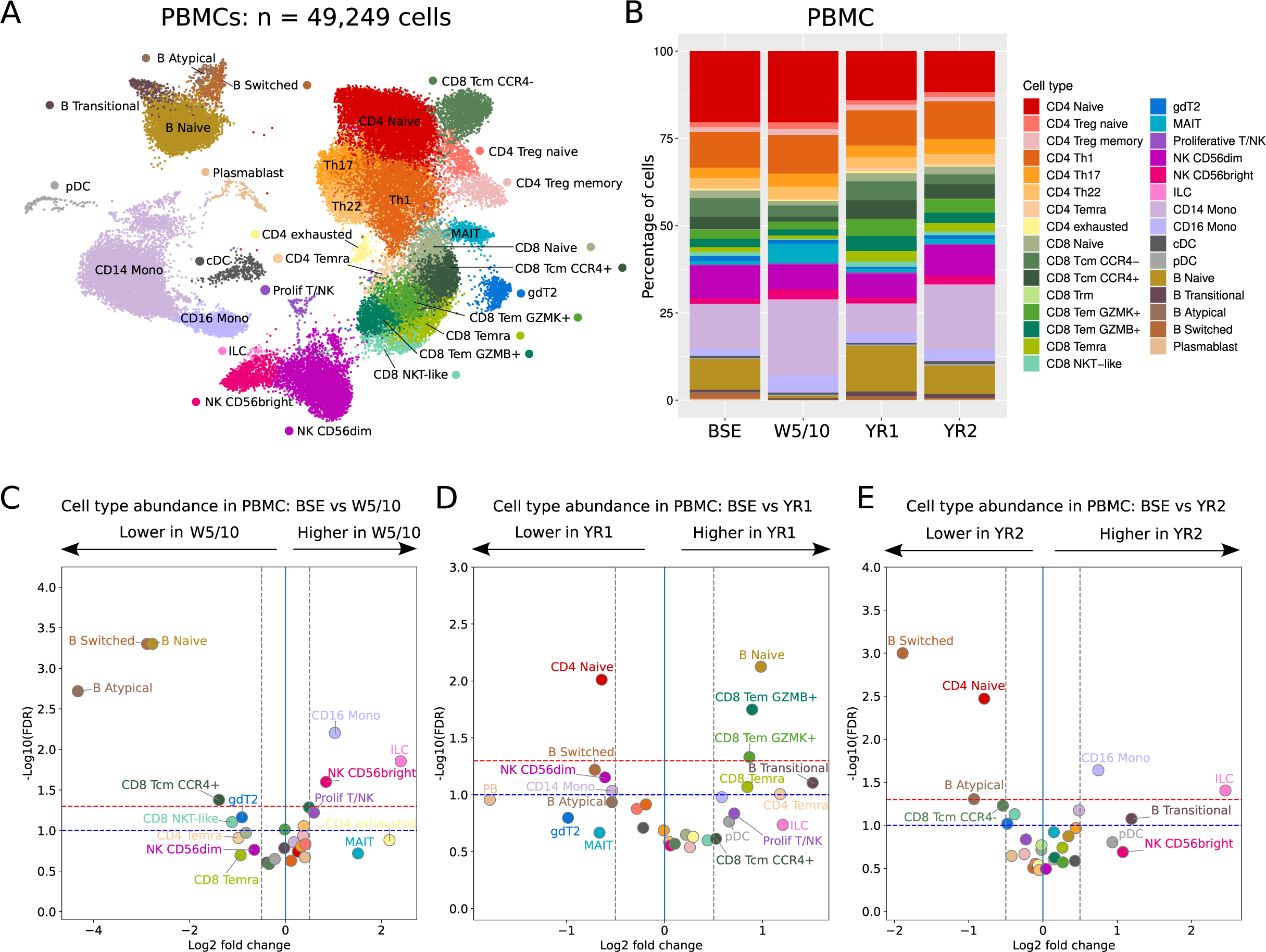
CladT-mediated changes in PBMC subpopulations at single cell resolution. (**A**) UMAP depicting PBMCs from all subjects. (**B**) Percentage of each PBMC cluster at four different timepoints throughout CladT treatment. (**C–E**) Volcano plot depicting differences in cluster abundance from PBMCs. Immune cell subset changes at BSE compared to W5/10 **(C)**, YR1 **(D)**, and YR2 **(E)** revealed by fold change (log2) versus p value (-log10) using sscomp analysis. The blue and red horizontal dashed lines indicate significance thresholds of 0.1 and 0.05, respectively.

While several subpopulations of B cells were initially depleted, blood T cells were less affected by CladT therapy. No significant changes in CD4 T cell subsets were found at early time points, although a trend toward significance was observed in the reduction of CD4 Temra cells at week 5-10. At year one post-CladT, naive CD4 T cells were significantly reduced in blood and remained depleted at year 2 (**Figure 2D, E**). Some subsets of CD8 T cells such as Tcm CCR4+ declined in proportion earlier (5/10 weeks) and were found to be persistently reduced at year 2 after CladT treatment (**Figure 2C–E**). Notable at one year post-CladT therapy was a significant elevation in Tem subsets of CD8 T cells (GZMB+ and GZMK+) compared with baseline, normalizing by year 2 (**Figure 2C–E**).

CladT-mediated effects on subpopulations of NK cells, innate lymphoid cells (ILCs), and monocytes were also observed in the blood. In particular, CD56^dim^ NK cells were noticeably reduced at year 1 after CladT therapy, approaching statistical significance. However, by year 2 CD56^dim^ NK cells were at levels similar to baseline. In contrast, CD56^bright^ NK cells, ILCs, and CD16 monocytes demonstrated consistently higher cell abundance compared to baseline at most time points after treatment (**Figure 2C–E**). Differential gene expression (DEG) analysis of PBMC subpopulations between baseline vs. weeks 5/10 and baseline vs. year 1 subgroups demonstrated major changes in the plasmablast cluster after CladT therapy (**Supplemental Figure 4A, B**). The downregulated genes expressed by plasmablasts following CladT therapy (Immunoglobulin heavy and light chain genes) potentially indicate a change in B cell clones. Overall, these data demonstrate a broad, early effect of CladT therapy on several immune cell populations in the blood, including T and B cell and myeloid subsets, with sustained changes in naive CD4 T cells as well as switched B cell subsets over time.

### Alterations in immune cell composition within the CSF after CladT treatment for relapsing MS

CladT-associated changes in the CSF of MS patients have not been extensively described, despite the CSF serving as a better reflection of the CNS compartment where MS lesions actually manifest. To address the mechanism by which CladT therapy for MS exerts effects within the CNS compartment, we analyzed a total of 11 CSF samples from seven MS patients at different time points before and after CladT treatment and analyzed the cell type composition captured by scRNA-seq (**Figure 1A**). We identified and subclustered 23,298 CSF cells resulting in 32 distinct subpopulations (**Figure 3A**). When examining the frequency of different B cell subsets we found, similar to blood, a reduction in naive B cells at weeks 5/10 followed by an increase in proportion at year 1 compared with baseline (**Figure 3B-D**). The changes observed in the CSF switched B cells paralleled those in blood, with a reduction at weeks 5/10 persisting as a statistically significant decrease at year 1 (**Figure 3C, D**). The frequency of CSF plasmablasts was elevated at week 5/10 post-CladT therapy but normalized to baseline at year 1, while a trend toward an increase in transitional B cell frequency after 1 year of CladT treatment was observed (**Figure 3B-D**).

**Figure 3.**
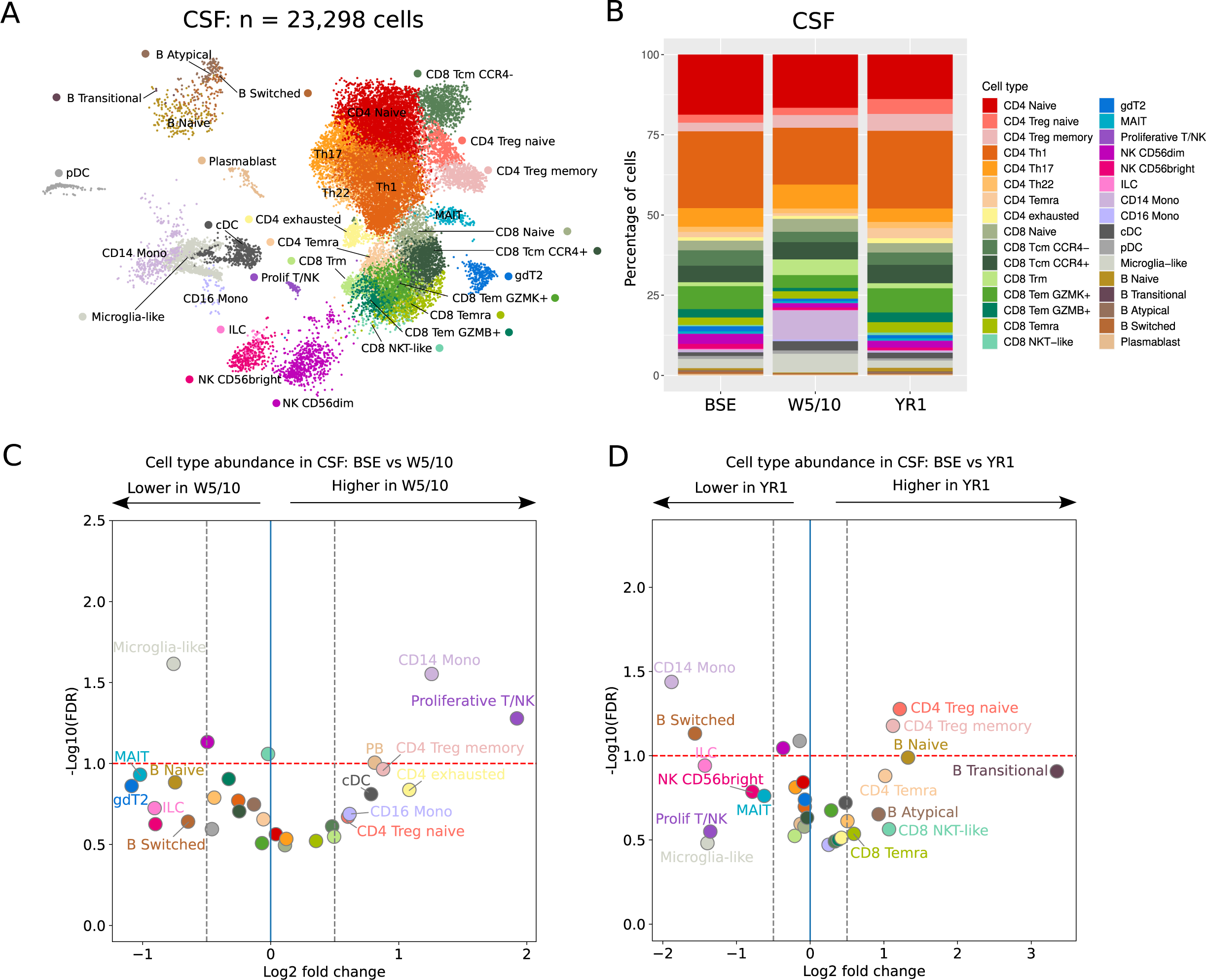
CladT-mediated changes in CSF subpopulations at single-cell resolution. (**A**) UMAP depicting CSF cells from all subjects. (**B**) Percentage of each CSF cluster at three different timepoints after CladT treatment. (**C, D**) Volcano plot depicting differences in CSF cluster abundance. BSE compared to W5/10 (C) and YR1 (D) as revealed by fold change (log2) versus p value (-log10) using the sscomp methodology. The red horizontal dashed line indicates significance threshold indicates a significance threshold of 0.1.

Examining CD4 T cell subsets in the CSF compartment, we observed striking CladT-associated remodeling in CD4 T cell subpopulations. In particular, an increase in the relative abundance of CSF CD4 T regulatory (Treg) memory cells was seen soon after CladT therapy with persistent elevation at year 1. The Treg naïve subpopulation was significantly elevated in the CSF one year following CladT therapy as well (**Figure 3B–D**). This was also accompanied by an elevation of the CD4 Temra subcluster at one-year post-CladT treatment, although this change did not reach statistical significance. In contrast to blood, we did not find any changes in the proportion of CSF CD4 naive T cells. While exhausted CD4 T cells were found to be elevated in the blood and CSF at weeks 5/10, this difference did not reach statistical significance. CD8 T cell subsets in the CSF were affected less by CladT therapy compared to blood; at weeks 5/10 there were no changes in the abundance of Tcm CCR4+, naive, and NKT-like CD8 T cells compared to baseline.

Among other cell types, we noticed changes in the innate lymphoid subset of immune cells within the CSF after CladT therapy. CSF microglia-like cells were drastically reduced early after CladT therapy (**Figure 3B–D**). In contrast, CD14+ monocytes were significantly more abundant in the CSF early after CladT therapy. However, after one year, CSF monocytes were found to be drastically reduced (**Figure 3B-D**). At year 1 post-CladT therapy, microglia-like cells and CD14 monocytes were also slightly less abundant than pre-treatment. Through DEG analysis of CSF subpopulations between baseline vs. week 5/10 and baseline vs. year 1, we noted remodeling of the plasmablast cluster. Similar to changes observed in the blood, transcriptional changes in plasmablasts were characterized by a lower diversity of immunoglobulin heavy and light chains after CladT treatment (**Supplemental Figure 4C, D**). In sum, CladT treatment resulted in parallel blood and CSF depletion of B switched memory cells along with the emergence of CSF-specific emergence of CD4 Treg naive and memory subsets that could play a protective role in MS pathogenesis by suppressing inflammation and autoreactive immune cells.

### CladT-mediated T cell clonal dynamics

With the extensive literature indicating an important role of clonally expanded autoreactive T cells in MS pathogenesis and progression^30,31^, we wanted to explore the dynamics of CD4 and CD8 T cell clonality in both blood and spinal fluid from MS patients throughout CladT therapy. Hence, we profiled TCR sequences using the 10x Genomics platform from 34 samples obtained from 13 MS subjects treated with CladT. After quality control, we obtained a total of 37,223 TCR sequences representing blood and CSF compartments combined from baseline, weeks 5/10, and years 1 and 2.

As expected, we observed substantially higher clonality in the CD8 T cell population compared to CD4 T cells from the entire object (**Figure 4A, Supplemental Figure 5A-D**). Clonality of CD8 T cells did not substantially differ between blood and CSF at baseline, while higher overall clonality of CD4 T cells in the CSF versus blood was seen before CladT therapy **(Supplemental Figure 5E, F)**. This was driven by significantly higher CSF clonality in CD4 Treg memory and CD4 exhausted subsets and with a trend toward significance in CD4 Th1 cells (**Supplemental Figure 5G**). Among CD8 T cell clusters, we also found CSF CD8 Tcm CCR4-to be the most consistently expanded compared to blood prior to treatment. These findings emphasized the unique immune microenvironment of the CSF in MS in relation to T cell repertoire diversity.

**Figure 4.**
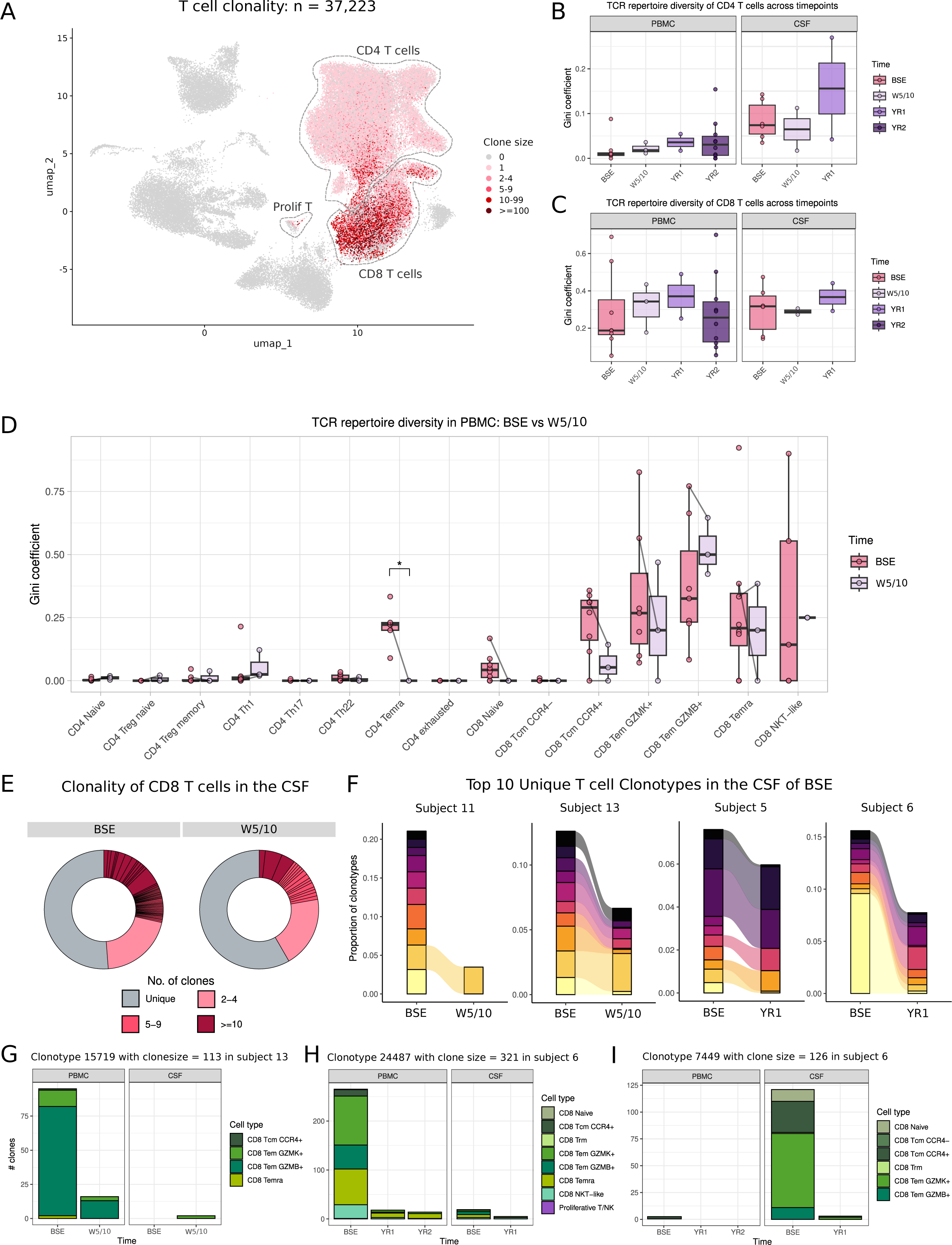
Clonal dynamics of T cells after CladT treatment of MS subjects. (**A**) UMAP depicting clone size of T cells. (**B, C**) TCR repertoire diversity of CD4 (**B**) and CD8 (**C**) T cells from PBMCs and CSF at BSE and WK5/10, YR1, and YR2 post-CladT treatment. (**D**) TCR repertoire diversity across subsets of T cells from PBMCs at BSE vs W5/10. (**E**) Donut plot of CD8 TCRαβ clonality from the CSF at BSE and W5/10 post-CladT treatment. Clones are colored based on their clone size. (**F**) The top 10 clonotypes from the CSF at BSE for subjects 5, 6, 11, and 13 as a relative proportion of clonotypes for corresponding BSE, W5/10 or YR1 samples. Each color represents a unique clonotype by subject. (**G–I**) Clonal dynamics and composition of representative clonotypes from PBMCs and CSF (absolute number). In **B–D,** boxplot whiskers indicate values within 1.5 × interquartile range (IQR) from either upper or lower hinge. Horizontal bars represent the median value. Significance for pairwise comparison between BSE and all other timepoints by post-hoc Tukey test with False Discovery Rate (FDR) correction: *p.adj < 0.05.

When we compared the overall TCR sequences from CD4 and CD8 T cells in blood and CSF after CladT treatment, no significant difference in clonality, as measured by Gini coefficient, was seen (**Figure 4B, C**). However, we observed a drastic depletion of CD4 Temra clones along with a trend for decreased clonality in CD8 naive and CD8 Tcm CCR4+ subpopulations in the blood at weeks 5/10 post-treatment (**Figure 4D**). In the CSF at weeks 5/10 following CladT treatment, large CD8 T cell clones (>10 cells per clone) decreased substantially (**Figure 4E**). No other major differences in T cell clonality following CladT treatment were noted (**Supplemental Figure 6A-D**).

We next examined the dynamics of the top 10 T cell clones from the CSF of the four MS subjects for whom we had samples at both baseline and a subsequent time point, comparing baseline to weeks 5/10 or year 1. In all four subjects, at either 5/10 weeks or one year of CladT treatment, we found a partial depletion of the most abundant clones (**Figure 4F**). Additionally, we found these clones were almost exclusively CD8 T cells, specifically CD8 effector memory subsets such as CD8 Tem GZMK+, CD8 Tem GZMB+, and CD8 Temra **(Figure 4G-I, Supplemental Figure 7A-C**). In general, these clonotypes showed a dramatic reduction in their clone sizes following CladT treatment in both blood and CSF, although in one case re-expansion was observed (**Figure 4G-I, Supplemental Figure 7A-C**). Overall, these results indicate that CladT reduces large T clones in blood and CSF of MS subjects.

Given the interest in EBV infection as contributor to MS pathophysiology^32^, we also queried the specificity of TCRs using the publicly available VDJdb database^33,34^. Overall, 5,141 T cells were predicted to have specificity to viral peptides, 758 of which were annotated to be EBV-specific (**Supplemental Figure 7D, E**). Notably, among CD4 T cell subpopulations, we observed that only CD4 Th1 and CD4 Temra from CSF, and not blood, carried the potential to recognize viruses (**Supplemental Figure 7F**). Conversely, CD8 T cells exhibited a greater abundance of purported virus-specific TCRs in the blood, including Tcm CCR4+ and CD8 Temra clusters. Overall, these differences in virus-specific capacity of CD4 and CD8 T cell clones highlight the importance of tissue microenvironment in T cell activation and expansion and the importance of examining both CSF and blood to understand T cell clone dynamics.

### B cell clonal evolution after CladT treatment

To examine B cell clonality, isotype usage, BCR maturation processes, and tissue-specific differences associated with CladT therapy, we submitted 30 samples from 12 MS subjects for single-cell BCR sequencing. After filtering, we obtained a total of 5,046 B cells that contained BCR sequences (**Figure 5A**). Interestingly, we observed a prevalence of medium size clones in the CSF at baseline but more hyperexpanded (>10 cells) B cells in the blood (**Supplemental Figure 8A**). Plasmablasts were found to be the most clonal subset in both blood and CSF, whereas atypical memory B cells trended toward tissue-specific clonality with expansion in the CSF (**Supplemental Figure 8B**). We did not find any significant changes in clonality within blood or CSF B cell subclusters after CladT treatment (**Supplemental Figure 8C–H**).

**Figure 5.**
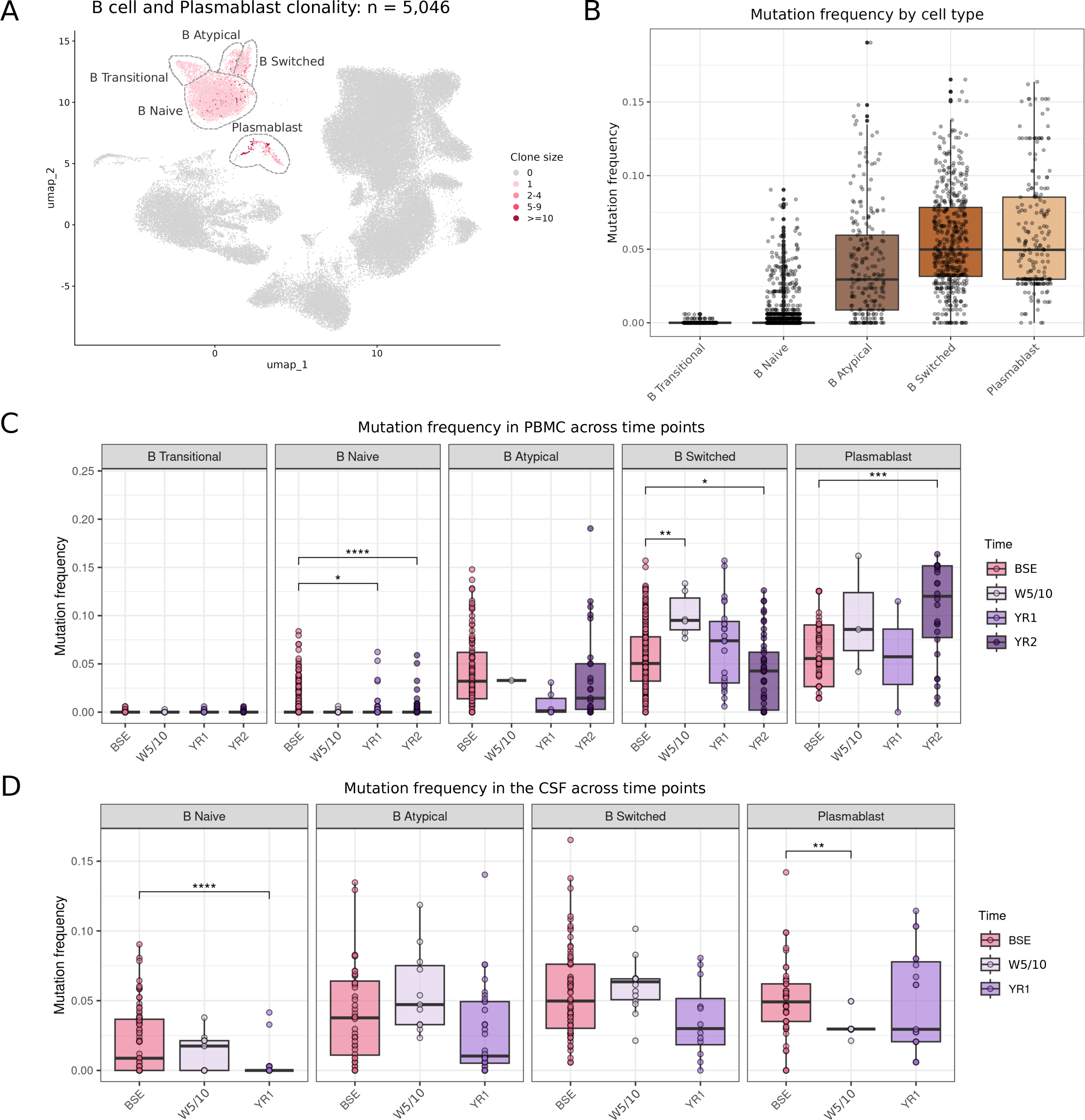
BCR mutational dynamics after CladT treatment. (**A**) UMAP depicting clone size of B cells. (**B**) BCR mutation frequency in B cell subsets from both PBMCs and CSF. (**C, D**) BCR mutation frequency in B cell subsets at BSE, WK5/10, YR1, and YR2 timepoints post-CladT therapy. PBMCs **(A)** (C) and CSF (**D**).

To assess changes in isotype usage, we calculated the proportions of various immunoglobulins (Ig) in each B cell subset and compartment (**Supplementary Figure 9**). As expected, transitional and naive B cells were almost exclusively IgM-expressing, whereas atypical B cells, switched B cells, and plasmablasts predominantly expressed IgG or IgA (**Supplemental Figure 9A, B**). Similar to a prior study^35^, we found a higher frequency of IgA1-expressing cells among plasmablasts in the blood, while plasmablasts expressing IgG1 or IgG2 were more abundant in the CSF **(Supplemental Figure 9C)**. Changes in Ig heavy chain usage following CladT therapy were modest and primarily observed in blood. In particular, atypical and switched B cell subpopulations from blood but not CSF exhibited a complete depletion of mature IgG-expressing cells at week 5-10, with the gradual recovery by year 1 and 2 **(Supplemental Figure 9D, E)**.

We next investigated the dynamics of somatic hypermutation (SHM) to better understand CladT-mediated effects on affinity maturation. As expected, we found low SHM frequencies in *IGHM* and *IGHD* and significantly greater rates of SHM in IgA and IgG isotypes (**Supplemental Figure 10A**). Moreover, when comparing the frequency of mutations in each isotype from blood and CSF, we noticed a statistically significant difference in the proportion of IgM and IgD chain mutations in B cells from the CSF (**Supplemental Figure 10B**). In line with a prior report^36^, transitional and naive B cells showed the lowest SHM frequencies, whereas atypical B cells, switched B cells, and plasmablasts acquired a much greater mutation frequency (**Figure 5B, Supplemental Figure 10C, D**). Among naive B cells defined similarly to Ramesh and colleagues^35^ by expression of genes such as *IL4RA* and *FCER2A* (and which almost exclusively express IgM, **Supplemental Figure 9B**), we found that the SHM frequency was significantly higher in the CSF compared to blood (**Supplemental Figure 10D**). A significant effect of CladT therapy was observed on the frequency of mutations in both compartments when examining specific B cell subsets. Specifically, despite low mutational frequency in naive B cells, an even lower SHM frequency was observed following CladT therapy in blood and CSF at year 1 (also seen at year 2 in blood; **Figure 5C, D**). In contrast, switched B cells had a higher frequency of accumulated mutations at weeks 5/10 (statistically significant in the blood), but returned to baseline levels by year 1 of therapy. Curiously, a statistically significant reduction at year 2 post-CladT therapy in SHM of switched B cells in blood was then observed compared to baseline, while mutational frequency in blood plasmablasts was increased relative to baseline **(Figure 5C)**.

To also determine the effect of CladT treatment on the depletion and maturation of individual B cell clones in the context of MS, we attempted to reconstruct B cell lineage trees starting with germline Ig sequences. In general, we found it difficult to identify clones present both before and after CladT therapy, particularly in both CSF and blood compartments. Nevertheless, subject 13 allowed us to reconstruct several clonal trees, three of which are shown in **Supplemental Figure 10E-G**. Clone 747 consisted of both naive and switched B cells that exhibited a low number of substitutions in the heavy chain of IgM (**Supplemental Figure 10E**). This clone persisted at week 5 only in the CSF following treatment with CladT. Another, clone 438 consisted predominantly of IgG-expressing plasmablasts which persisted and potentially expanded in the CSF after CladT-therapy (**Supplemental Figure 10F**). Clone 1941 consisted of plasmablasts that exhibited a much higher mutational load vs the germline sequence in the heavy chain of IgA. This clone was only found at baseline in the blood and not following treatment with CladT (**Supplemental Figure 10G**). In summary, our data show that CladT alters the composition of B cell subsets, rapidly depleting most of the memory B and plasmablast clones in the blood out of proportion to the CSF.

## Discussion

CladT is a highly effective DMT for the treatment of MS^9,12^. Mechanistically, CladT not only inhibits DNA synthesis and repair in dividing lymphocytes but reduces energy production in quiescent lymphocytes, leading to selective depletion of B and T cells. While CladT has been shown to cross the blood brain barrier, with a CSF/plasma concentration ratio of approximately 0.25 in the context of cancer^37^, a comprehensive understanding of the effect of CladT on immune cells in the CNS compartment and the cellular dynamics that accompany amelioration of MS following CladT therapy is needed.

Using scRNA-seq, we identified major effects of CladT on several of the 32 transcriptionally distinct immune subpopulations. Not only did we observe early changes in multiple populations of immune cells following CladT therapy, sustained alterations in specific lymphocyte subsets were detected by transcriptomic profiles of both the CSF and blood. Following CladT therapy, we found a long-lasting increase in relative proportion of Treg naive and Treg memory subsets in the CSF. An obvious potential benefit of Treg relative accumulation in the CNS compartment following CladT therapy stems from their capacity to suppress autoreactive^38^ and proinflammatory CD4 Th1 and Th17 T cells^39^, along with their ability to promote tissue repair and regeneration by modulating stem cell differentiation^40,41^. Longitudinal analysis of patients treated with CladT demonstrated an even greater expansion of some lymphocytes within the blood at year 2 compared with the first year following CladT therapy, including some regulatory subsets^42^. Hence, persistence of Treg subsets in the CSF at later times would not be unreasonable and could exert lasting benefits associated with the durable effect of CladT therapy for MS^43–45^. We hypothesize that accumulation of regulatory CD4 T cell subsets in the CSF after CladT treatment is based on a unique CNS compartment cellular repopulation that differs from blood rather than from transcriptional changes in existing T cell subsets induced by CladT. Unlike in the blood, we observed a preservation of CD4 naive T cells in the CSF; this population could serve as progenitors for CD4 Treg subsets specifically in the CSF. Further, our analysis did not find any significant phenotypic changes in Treg subsets, such as cytokine or chemokine expression, after CladT therapy (data not shown).

We have also examined the immunological consequences of CladT in MS through assessment of lymphocyte receptor sequences. While we did not observe a significant change in overall clonality of CD4 and CD8 T cells in either blood or CSF compartment, a specific reduction of hyperexpanded CD8 T cell clones predominantly in the CSF beginning at weeks 5/10 after CladT therapy was observed. These CD8 T cell clones showed a pronounced migration pattern from blood to CSF and were mainly composed of memory subsets, including Tem GZMB+, Tem GZMK+ and Temra. Mutation frequency analysis of BCRs revealed a substantial decrease in the number of mutations in naive B cells after 1 year (blood and CSF) and even 2 years (blood) following CladT treatment. Similar parallels between the blood and CSF were found in the reduction of mutation frequency in memory switched B cells, along with a similar trend in the atypical memory B subset. With this in mind, the selective depletion of certain B cell subsets could reshape the clonality of B cells persisting in both compartments by generating a pool of fresh B cells emergent in the blood as well as CSF. In particular, after drastic reductions of naive and memory B cell subsets, a new pool of B cells with less mature antibodies could replenish those more antigen-experienced B cells. BCR repertoire analysis revealed that CladT treatment reduces the number of clonally-specific mature memory B cells in the blood at early time points. Importantly, in contrast to blood, the abundance of atypical memory B cells in the CSF were only slightly affected early after CladT therapy, suggesting its unique effects on spinal fluid cells. This exemplifies the subtle but important compartment-specific differences in the effect of CladT therapy on B cell subsets despite similar dynamics in cell proportions.

Several limitations of this study should be considered. First, larger subject cohorts will be necessary to validate our findings of CladT-mediated dynamics within the CSF compartment. Second, the low number of cells available for study prevented more rigorous computational analysis, impacting, for example, defining the clonality of memory B cells in the blood at weeks 5/10 post-CladT treatment. This is likely due to both selective CladT effect on B memory subpopulations and a small sample size for that early time point. Third, there might be further immunological dynamics in the CSF beyond 1 year from initiation of CladT treatment. Thus, further studies extending the observation period of CSF samples to better understand the long-term effect of CladT therapy on the CNS would be valuable. Finally, we did not directly correlate the outcome measures employed by CLOCK-MS, such as disability measures or MRI lesion burden, with the kinetics of blood and CSF subsets. Determining an association between disease activity and changes in cellular composition, in particular within the CSF, will be one important avenue to pursue.

With increasing realization of the importance of compartmentalized cellular inflammation in MS, we have utilized scRNA-seq to explore the effects of CladT therapy on the CSF of patients with relapsing MS. The recent characterization of CSF immune cells by scRNA-seq in MS^19,35^ and other neurologic diseases^46,47^ highlight not only the powerful utility of transcriptomic profiling through RNA sequencing but also the feasibility of these experiments. Overall, our study reveals similar long-lasting depletion of B memory subpopulations and unique repopulation dynamics of naive B cells in the blood and CSF upon CladT therapy. Furthermore, we have identified sustained CSF-specific emergence of CD4 Treg naive and memory subsets early on after CladT therapy which could serve as one mechanism by which CladT exerts durable benefit in MS. Extending these observations by combining this scRNA-seq data with ongoing and future studies on the CSF of MS patients treated with CladT and other DMTs will serve to uncover mechanisms of therapeutic efficacy as well as pathophysiology of MS.

## Methods

### Subjects

Subject information is described in greater detail in the Supplemental Methods section and Table 1. Additional details regarding the enrollment criteria for CLOCK-MS can be found at https://clinicaltrials.gov/ct2/show/NCT03963375.

**Table 1.** Characteristics of study subjects.

|  | **Total (N=13)** |
| --- | --- |
| **Age (years)** | 48.8 ± 11.1 |
| **Disease duration from symptom onset (years)** | 14.0 [8.0,22.0] |
| **Disease duration from diagnosis (years)** | 13.0 [7.0,21.0] |
| **Gender n (%)** |  |
| . Male | 5 (38.5) |
| . Female | 8 (61.5) |
| **Race** |  |
| . White or Caucasian | 12 (92.3) |
| . Black or African American | 1 (7.7) |
| **Ethnicity** |  |
| . Not Hispanic or Latino | 13 (100.0) |
| **MS Type** |  |
| . RRMS | 12 (92.3) |
| . SPMS | 1 (7.7) |
| **EDSS** | 4.0 [1.5,4.0] |
| **Disease Activity** | 13 (100.0) |
| . Clinical Relapse | 9 (69.2) |
| . New Gadolinium (Gd) Enhancing Lesion | 5 (38.5) |
| . New T2 Weighted Lesion | 7 (53.8) |
| . Enlarged T2 Weighted Lesion | 2(15.4) |
| **Received Prior DMT** | 12 (92.3) |

Values presented as Mean ± SD, Median [P25, P75], Median (min, max) or N (column %).

### Sample preparation

The volume CSF samples varied between 5-30 mL, with a total number of 10,000 – 100,000 cells obtained per sample. Details regarding protocols involving sample processing can be found in the supplemental methods.

### Single-cell RNA-seq and analysis

CSF-cells and PBMCs were loaded into the 10x Genomics Chromium Controller for droplet-encapsulation, according to the manufacturer’s instructions. For sequencing the scRNA libraries were sequenced on an Illumina NextSeq or HiSeq 4000 to a minimum sequencing depth of 25,000 reads per cell. Details of sequencing data analyses can be found in the supplemental methods.

## Author Contributions

Roman A. Smirnov: Study design; data acquisition and analysis; drafting of the text and figures; revision of the manuscript for intellectual content.

Claudia Cantoni: Study design; data acquisition and analysis; drafting of the text and figures; revision of the manuscript for intellectual content.

Michael A. Paley: Data analysis; drafting of the text and figures; revision of the manuscript for intellectual content.

Kenneth Lee: Data acquisition and analysis; revision of the manuscript for intellectual content. Robert C. Axtell: Study concept and design; revision of the manuscript for intellectual content. Amber Salter: Study concept and design; data analysis; revision of the manuscript for intellectual content.

Samantha Lancia: Data analysis.

Brooke Hayward: Study concept and design; revision of the manuscript for intellectual content. Julie Korich: Study concept and design; revision of the manuscript for intellectual content.

Lori A. Lebson: Study concept and design; revision of the manuscript for intellectual content. Gabriel Pardo: Study concept and design; revision of the manuscript for intellectual content. Olaf Stuve: Study concept and design; revision of the manuscript for intellectual content.

Amit Bar-Or: Study concept and design; revision of the manuscript for intellectual content.

Maxim N. Artyomov: Study design; data analysis; revision of the manuscript for intellectual content. Anne H. Cross: Study concept and design; data acquisition and analysis; drafting of the text and figures; revision of the manuscript for intellectual content.

Brian T. Edelson: Study design; data analysis; drafting of the text and figures; revision of the manuscript for intellectual content.

Gregory F. Wu: Study concept and design; data acquisition and analysis; drafting of the text and figures; revision of the manuscript for intellectual content.

## Data Availability

All data produced in the present study are available upon reasonable request to the corresponding author.

## Acknowledgements

We would like to thank Isabel Risch for the draft of graphical illustrations in BioRender (Created in BioRender. Wu, G. (2025) https://BioRender.com/h63t991)

## Supplemental material

**Supplemental Figure 1.**
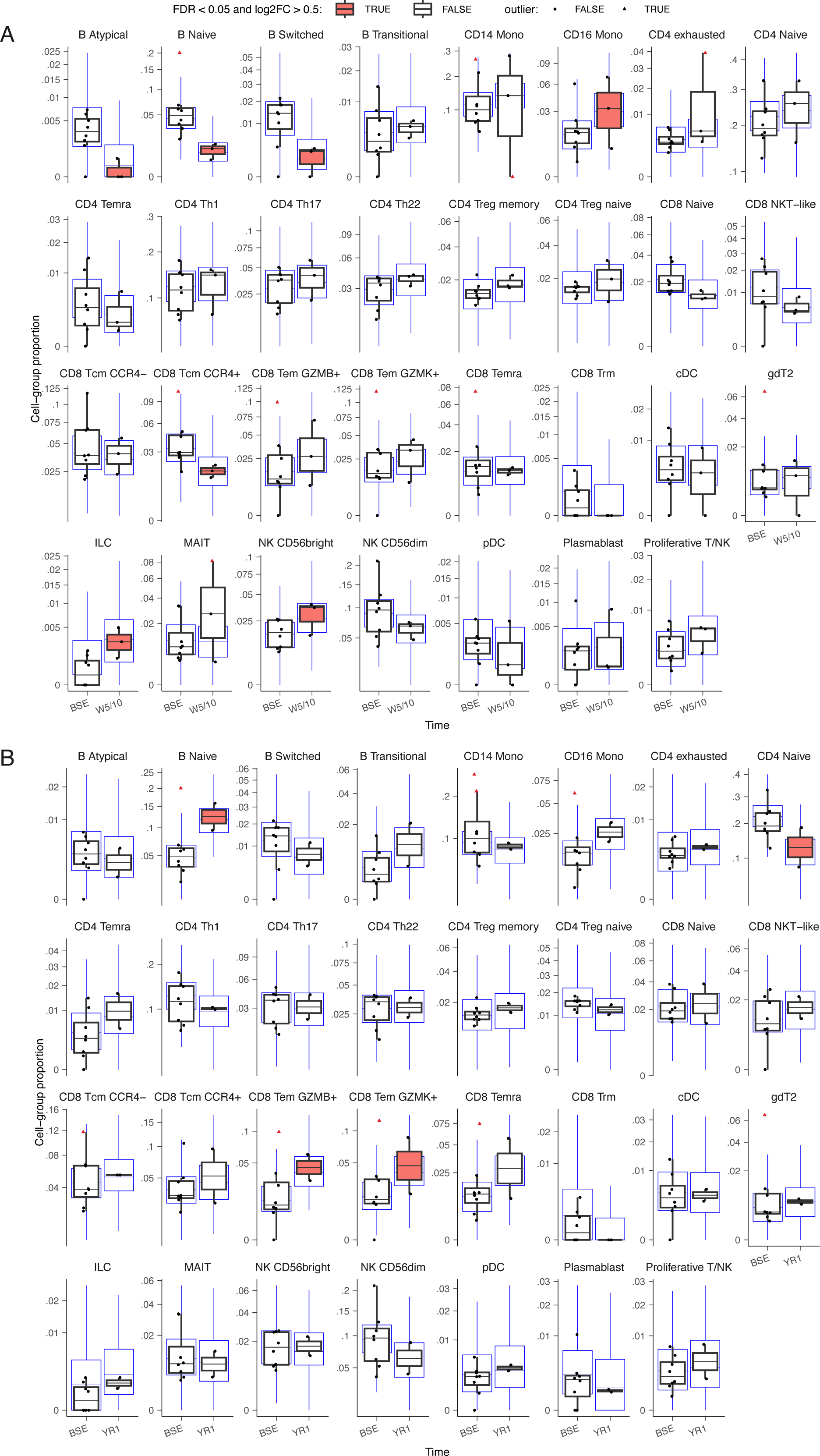
Differential composition analysis of cell type frequencies in PBMCs between baseline and post-CladT treatment. Box plots depicting differences in cluster proportions in PBMCs at BSE compared to (A) W5/10, and (B) YR1. Statistical significance, considered for an adjusted p value (FDR) < 0.05 and log2FC > 0.5 and highlighted in red, was calculated using sccomp. Error bars indicate 95% credible intervals with center lines representing medians. Blue outlines represent the posterior predictive check, which consists of a simulation from the fitted model; the overlap of simulated proportions with observed data was used as validation for the adequacy of the model. Red triangles represent predicted outliers.

**Supplemental Figure 2.**
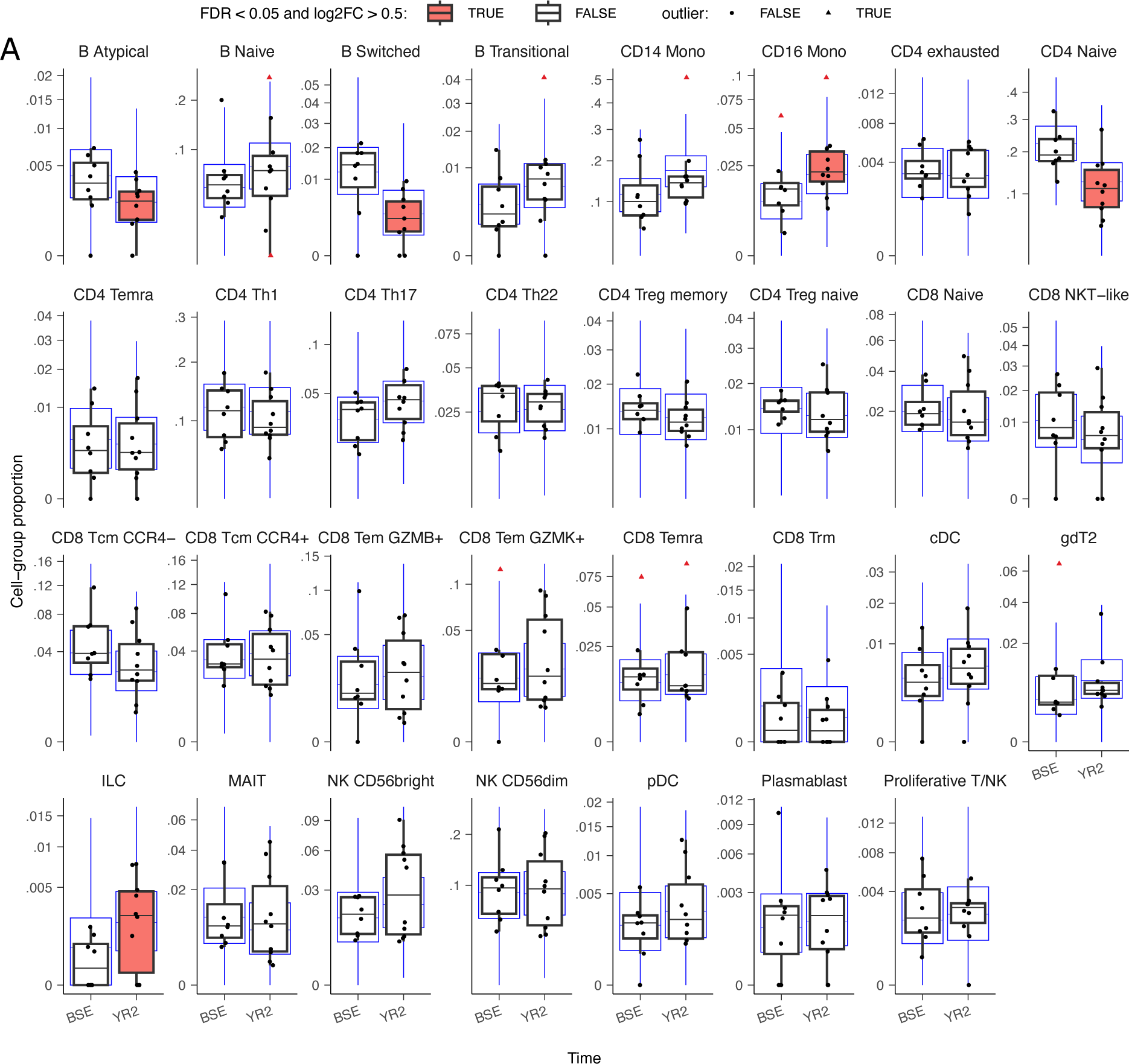
Differential composition analysis of cell type frequencies in PBMCs between baseline and YR2 post-CladT treatment. Box plots depicting differences in PBMC cluster proportions at BSE compared to YR2. Statistical significance, considered for an adjusted p value (FDR) < 0.1 and log2FC > 0.5 and is highlighted in red, was calculated using sccomp. Error bars indicate 95% credible intervals with center lines representing medians. Blue outlines represent the posterior predictive check, which consists of a simulation from the fitted model; the overlap of simulated proportions with observed data was used as validation for the adequacy of the model. Red triangles represent predicted outliers.

**Supplemental Figure 3.**
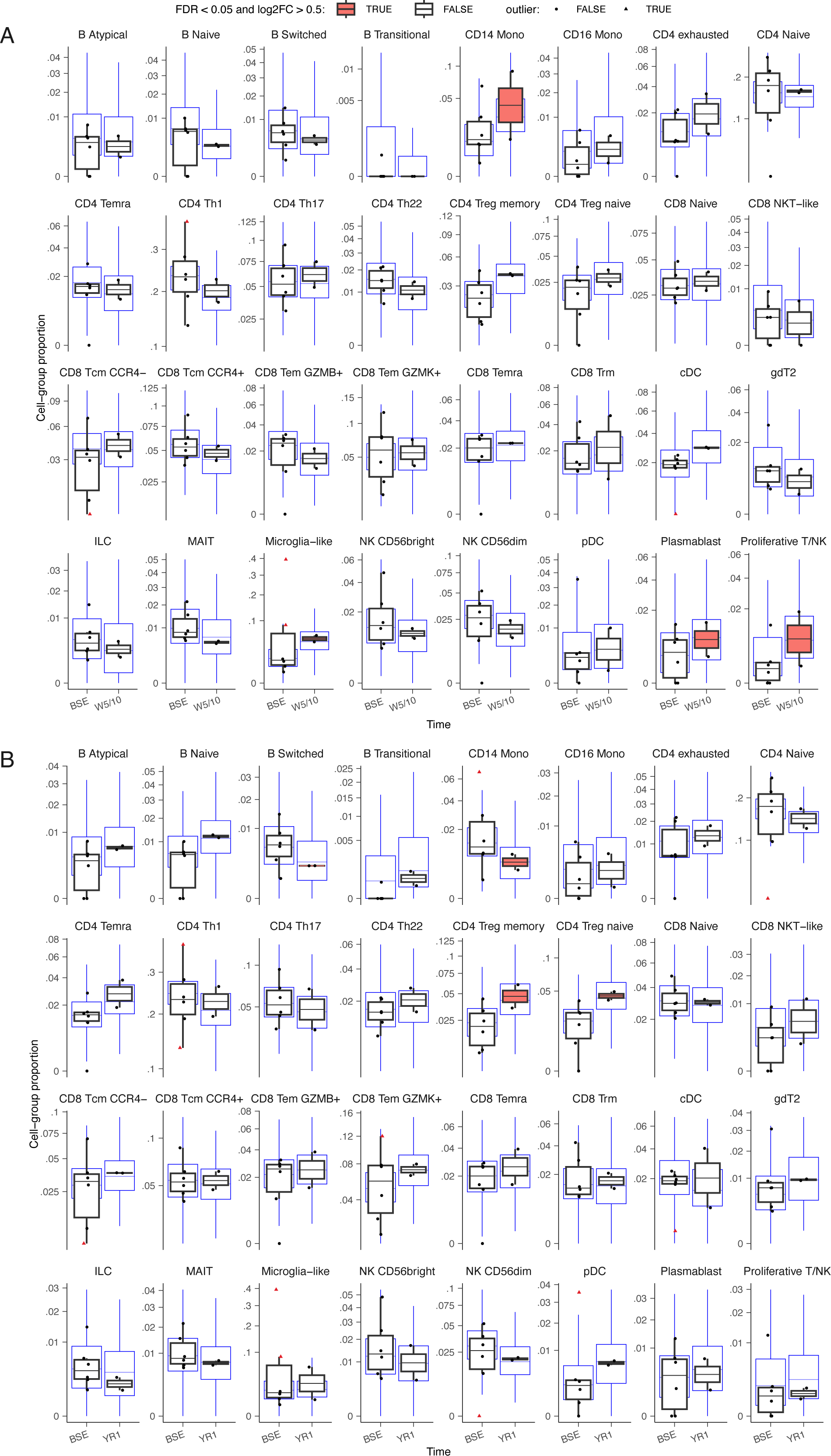
Differential composition analysis of CSF cell type frequencies between baseline and post-CladT treatment. Box plots depicting differences in cluster proportions in the CSF at BSE compared to (A) W5/10, and (B) YR1. model. Statistical significance, considered for an adjusted p value (FDR) < 0.1 and log2FC > 0.5 and is highlighted in red, was calculated using sccomp. Error bars indicate 95% credible intervals with center lines representing medians. Blue outlines represent the posterior predictive check, which consists of a simulation from the fitted model; the overlap of simulated proportions with observed data was used as validation for the adequacy of the model. Red triangles represent predicted outliers.

**Supplemental Figure 4.**
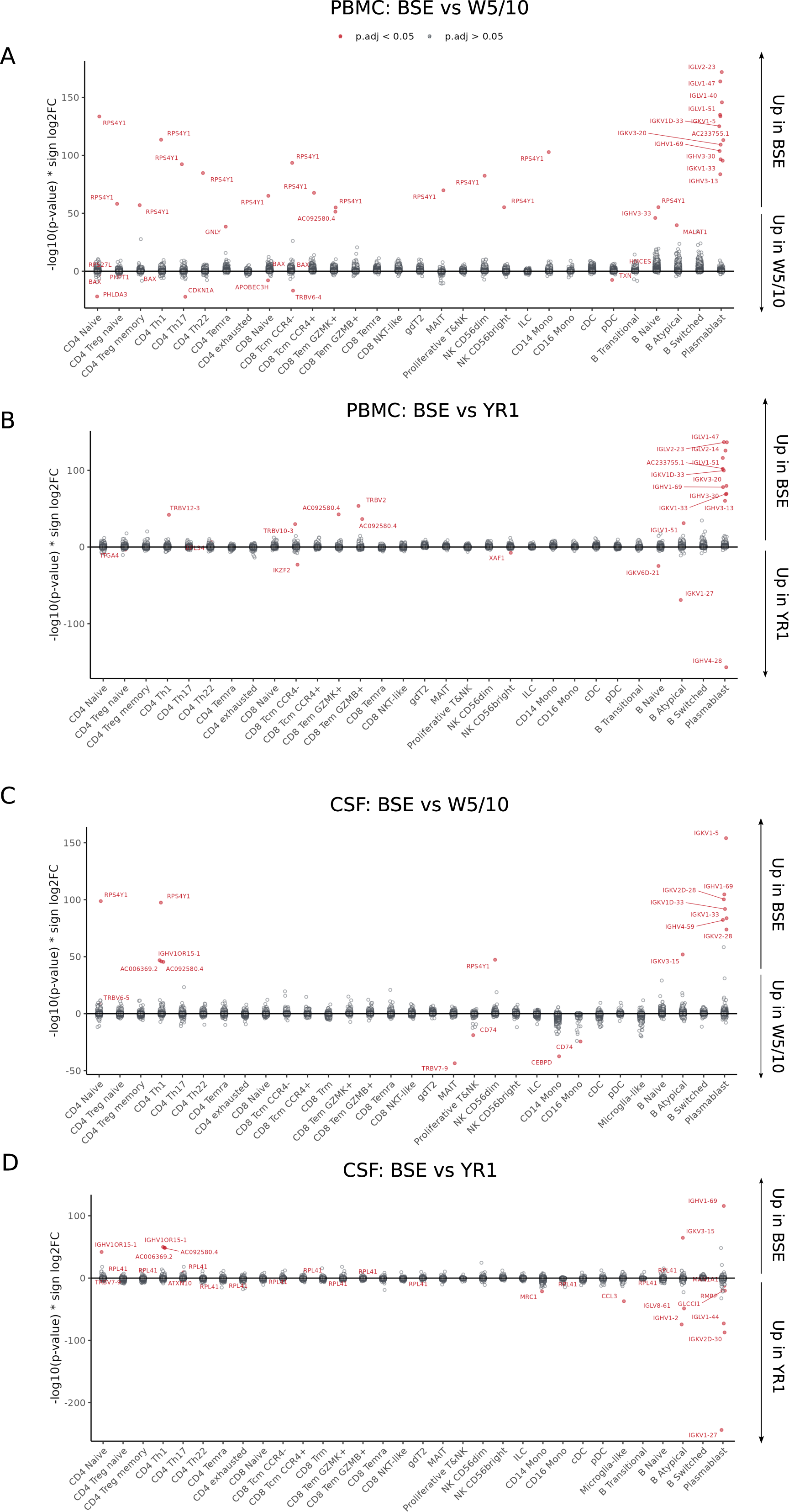
Differential expression analysis of all cell types after CladT treatment. Results of differential expression analysis between BSE and W5/10 (**A**) or YR1 (**B**) from PBMC, as well as BSE and W5/10 (**C**) or YR1 (**D**) from CSF across all subsets of cells.

**Supplemental Figure 5.**
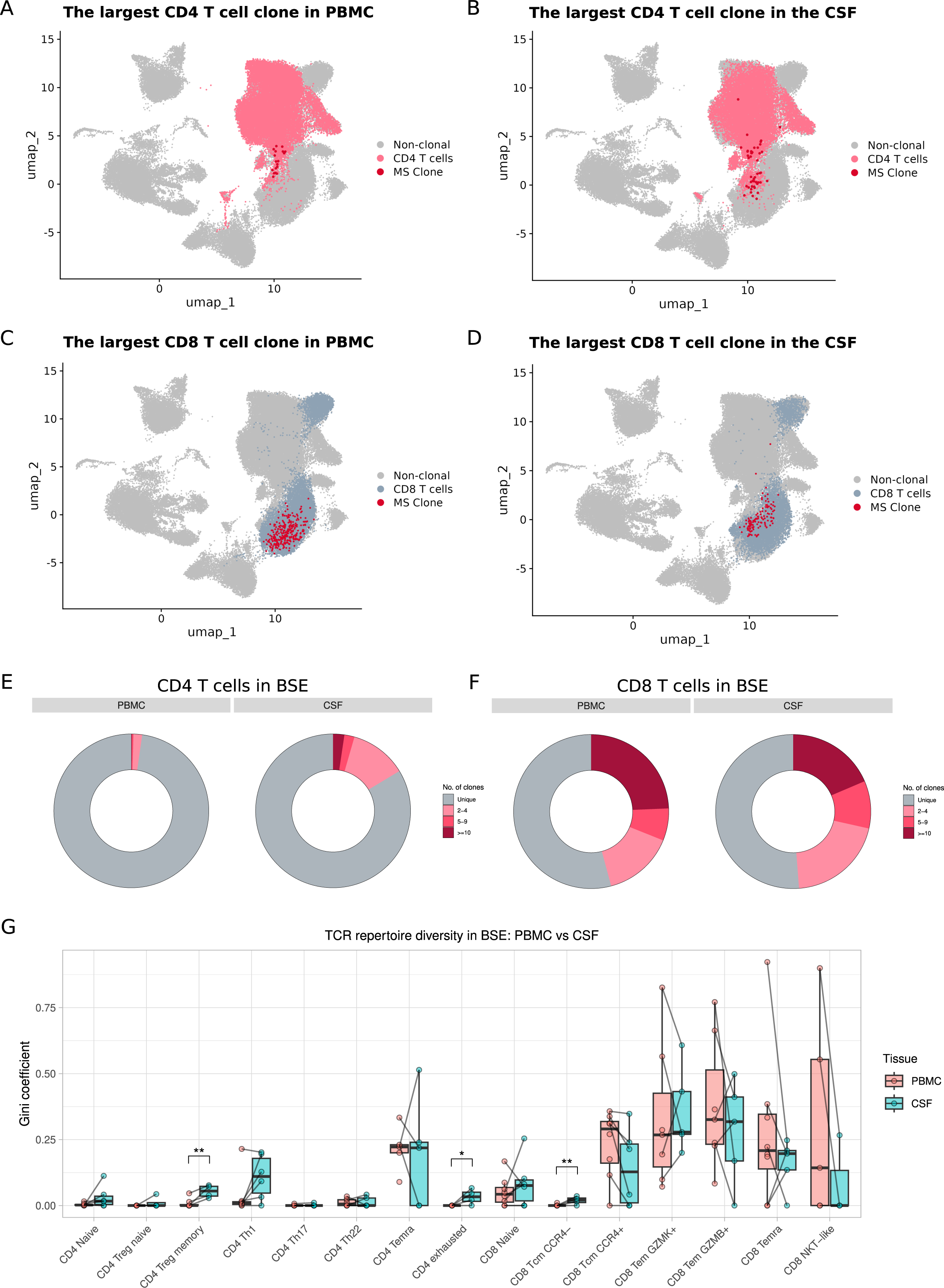
Compartment-specific differences in CD4 and CD8 T cell clonality between PBMCs and CSF across all time points. (**A–D**) Largest individual CD4 and CD8 T cell clones from PBMCs and CSF. Donut plots of CD4 (**E**) and CD8 (**F**) TCRαβ clonality from PBMCs and CSF at BSE. Clones are colored based on their clone size. (**G**) TCR repertoire diversity across subsets of T cells at BSE from PBMCs (red) and CSF (teal). In **G,** boxplot lower and upper hinges represent the 25th and 75th percentiles. Whiskers indicate values within 1.5 × IQR from either upper or lower hinge. Horizontal bars represent the median value. Significance on boxplots for pairwise comparison between HC and all other disease group by post-hoc Dunn test with False Discovery Rate (FDR) correction: *p.adj < 0.05, **p.adj < 0.01. These conventions are used for all subsequent Supplemental figures.

**Supplemental Figure 6.**
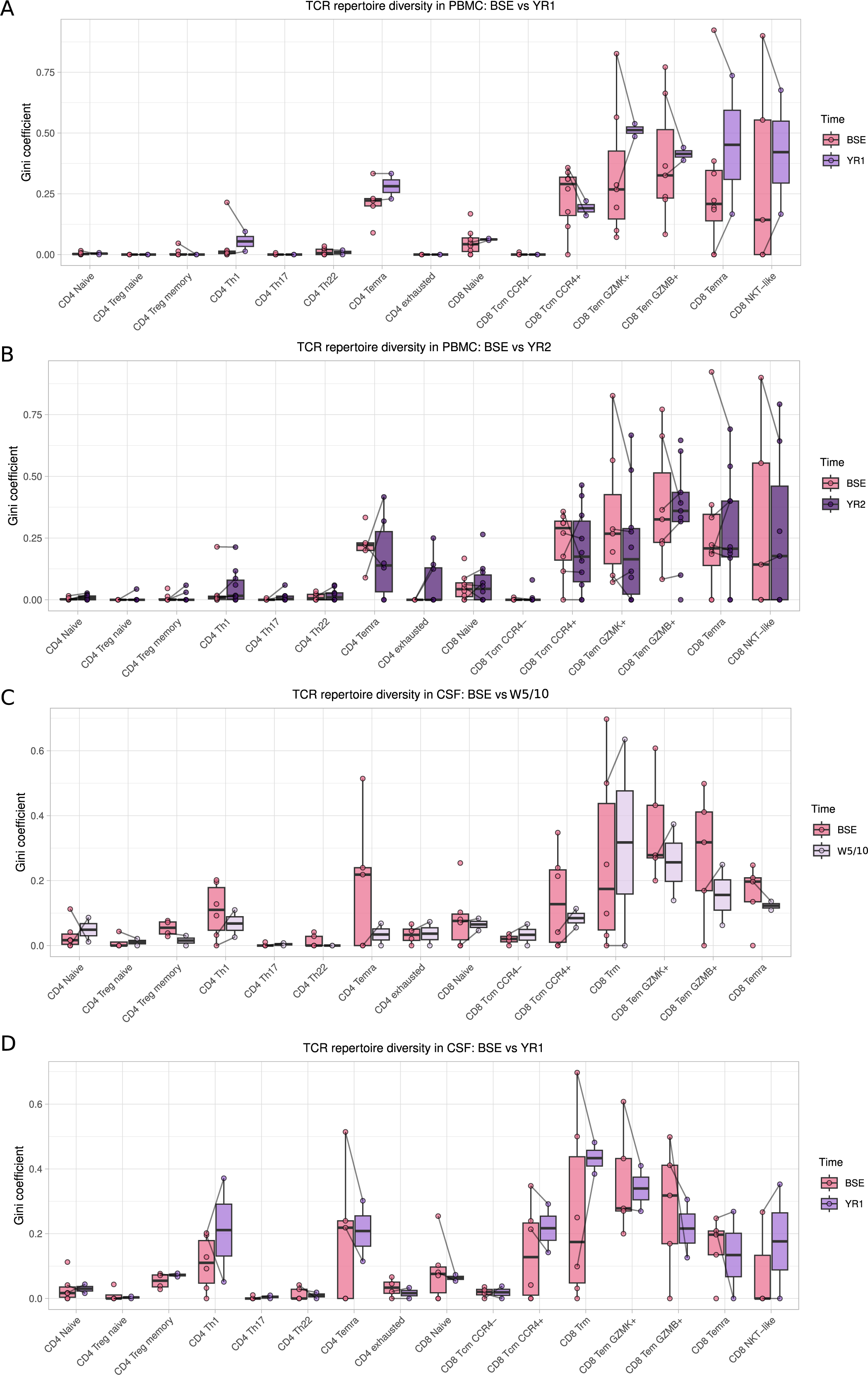
TCR repertoire diversity across all T cell subsets in PBMCs and CSF following CladT treatment. TCR clonality across subsets of T cells from PBMCs at (**A**) BSE versus YR1, (**B**) BSE versus YR2, as well as (**C**) from the CSF at BSE versus W5/10 and (**D**) BSE versus YR1.

**Supplemental Figure 7.**
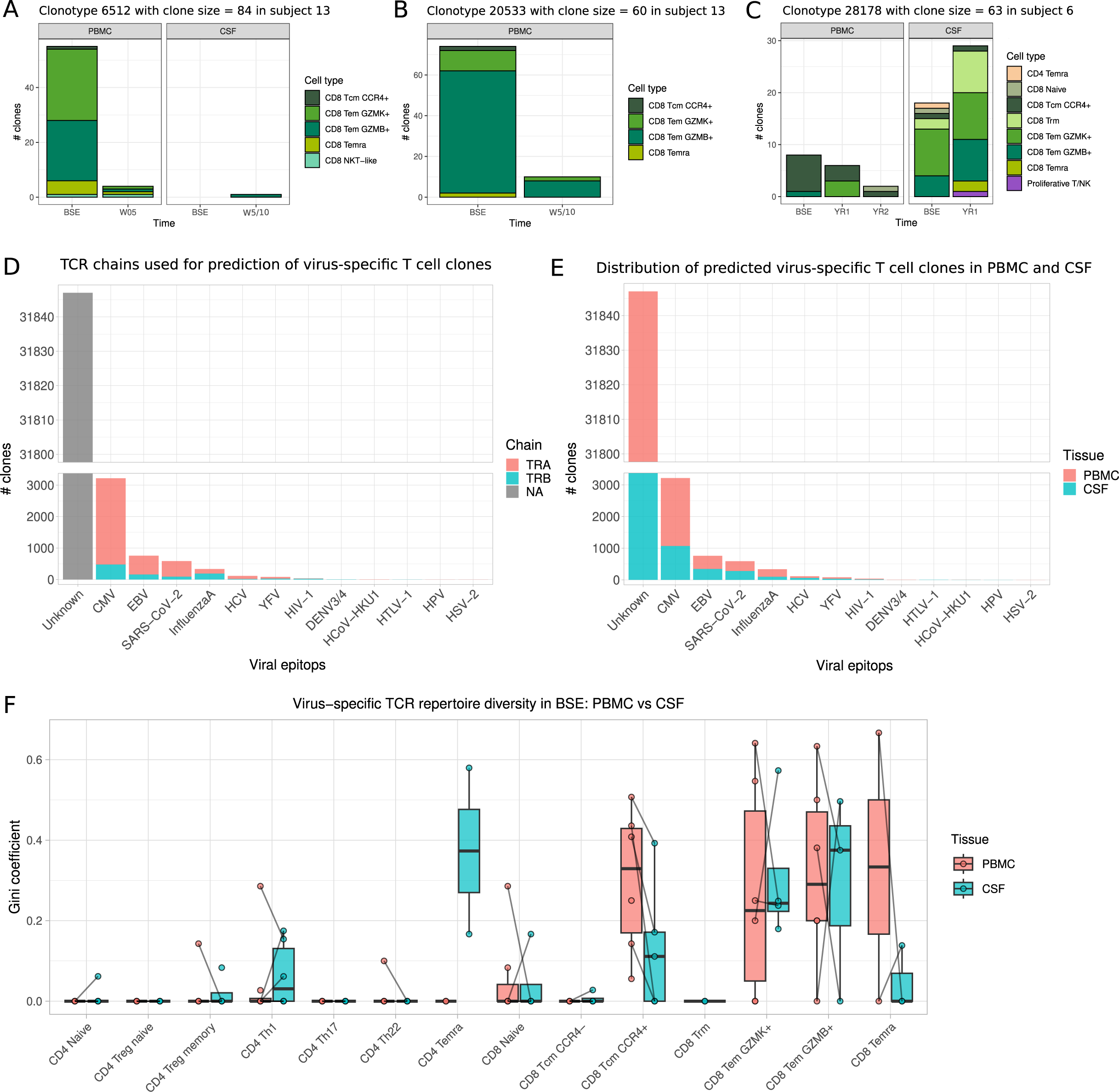
Differences in clonality of T cells from PBMCs and CSF with potential specificity to viruses. (**A–C**) Clonal dynamics and cell type composition of three top clonotypes from PBMCs and CSF. (**D**) The number of T cell clones predicted to have TCR specificities to viral antigens based on either alpha (red) or beta (teal) chain sequence and (**E**) their tissue-specific derivation. (**F**) Potential virus-specific clones from different T cell subsets within PBMCs (red) and CSF (teal).

**Supplemental Figure 8.**
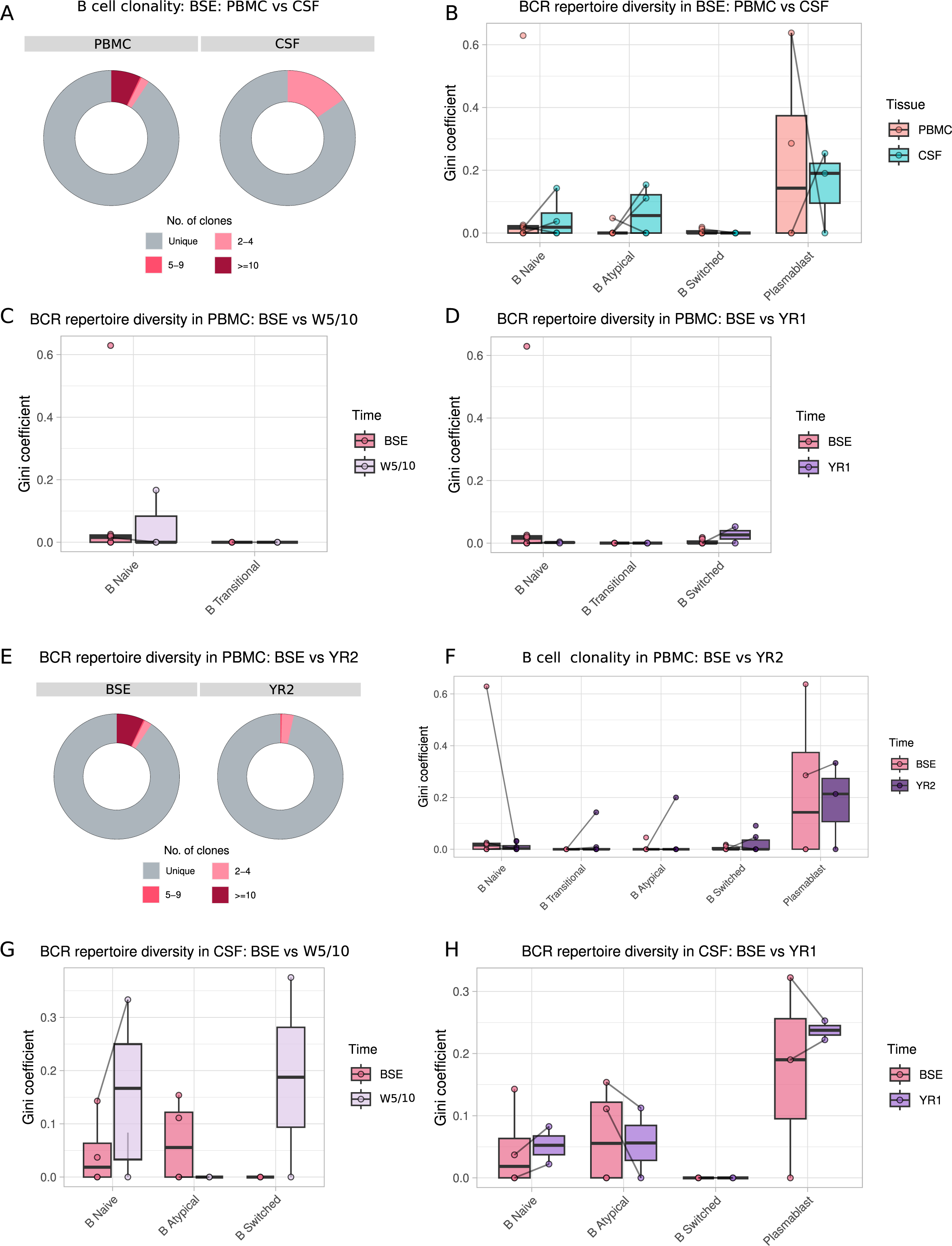
CladT-mediated changes in clonality of B cells from PBMCs and CSF. **(A)** Donut plot of B cell clonality from PBMCs and CSF at BSE. Clones are colored based on their clone size. (**B**) BCR repertoire diversity across subsets of B cells at BSE from PBMCs (red) and CSF (teal). BCR clonality across subsets of B cells from PBMCs at BSE versus W5/10 (**C**) and BSE versus YR1 (**D**). (**E**) Donut plot of B cell clonality from PBMCs at BSE versus YR2. (**F**) BCR clonality across subsets of B cells from PBMCs at BSE vs YR2. BCR clonality across subsets of B cells from CSF at BSE vs W5/10 (**G**) and BSE vs YR1 (**H**).

**Supplemental Figure 9.**
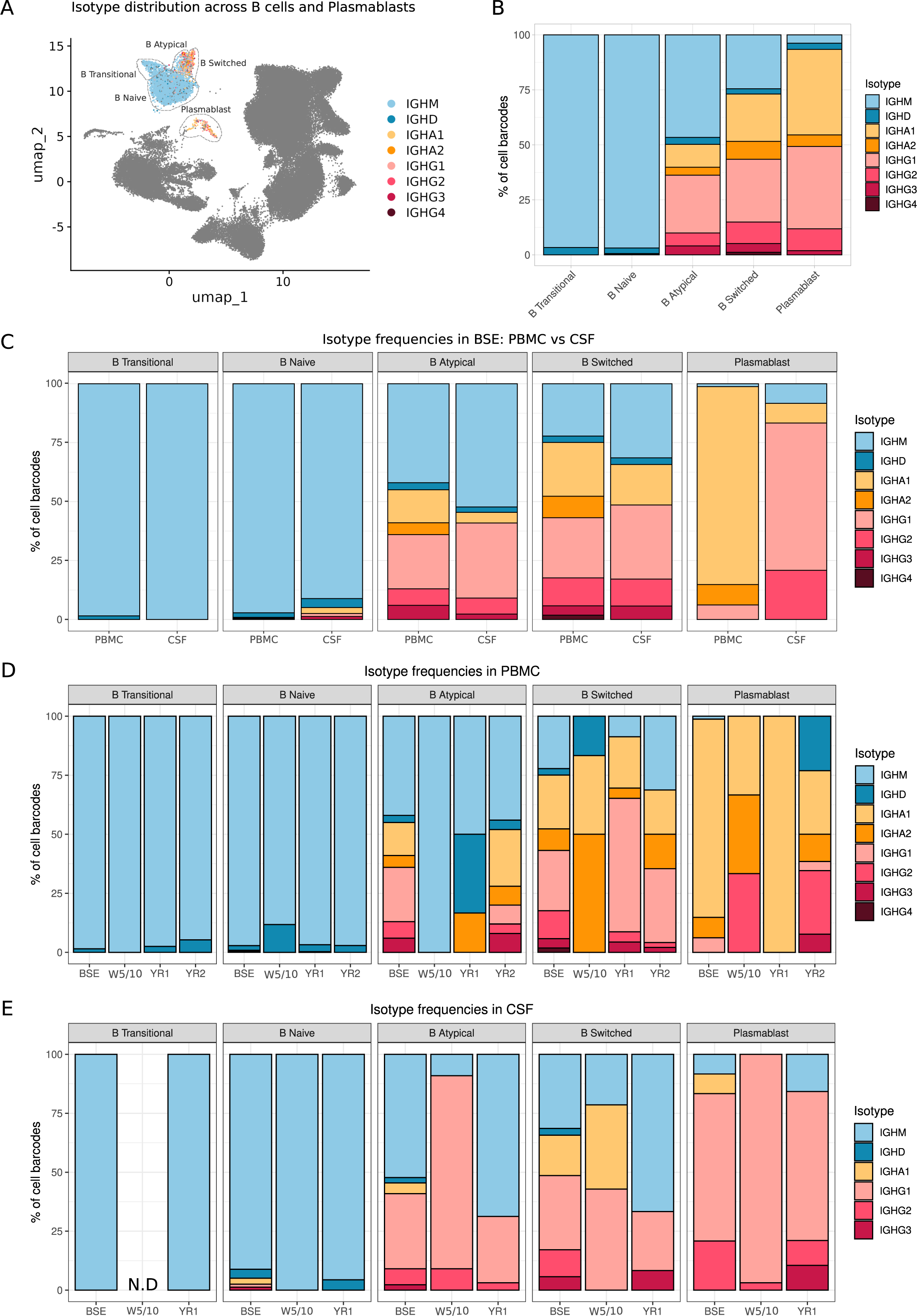
Compartment- and CladT-associated changes in antibody class usage across B cells and plasmablasts. (**A**) UMAP of different isotype usage in B cell and plasmablast clusters. (**B**) Relative frequencies of isotype usage by B cell subset. (**C**) Relative frequencies of isotype usage by B cell subset in both PBMCs and CSF at BSE. (**D, E**) Isotype usage across B cell and plasmablast clusters at different timepoints following CladT treatment from (**D**) PBMCs and (**E**) CSF.

**Supplemental Figure 10.**
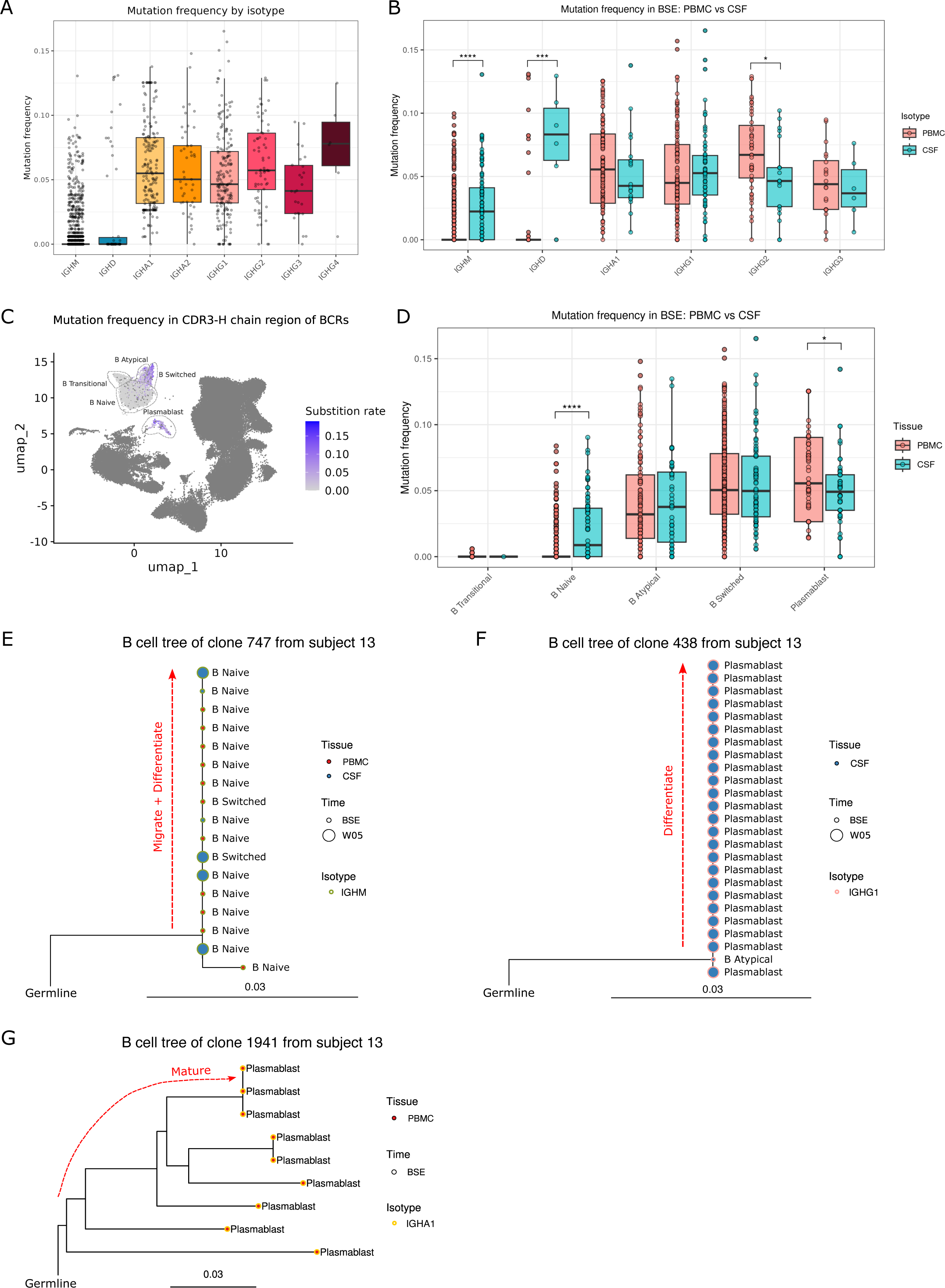
Compartment-specific differences in SHM across B cell subsets and plasmablasts. (**A**) Overall mutation frequency by antibody isotype. (**B**) Mutation frequency in antibody isotypes at BSE from PBMCs (red) and CSF (teal). (**C**) UMAP of mutation frequencies in CDR3-H chain region of paired BCRs. (**D**) Mutation frequency across subsets of B cells and plasmablasts at BSE from PBMCs (red) and CSF (teal). (**E–G**) Representative lineage trees of B cell clones from PBMC and CSF compartments of subject 13. Lineage tree topologies and branch lengths were estimated using standard maximum likelihood, with edge lengths representing the expected number of nucleotide substitutions per site as estimated using dnaml. Tips are colored by tissue type, and each tip’s border is colored by the antibody isotype.

**Table S1.** Sequencing samples.

| # | Subject ID | MGI folder ID | Tissue | Time-point | Sequencing | VdJ - BCR | VdJ - TCR |
| --- | --- | --- | --- | --- | --- | --- | --- |
| 1 | 1307_001_YR2_C | Wu_vdj_multi_MGI2375_10X | CSF | Year 2 | 5' seq | yes | yes |
| 2 | 1307_001_YR2_P | Wu_vdj_multi_MGI2375_10X | PBMC | Year 2 | 5' seq | yes | yes |
| 3 | 1307_002_YR2_P | Wu_vdj_multi_MGI2590_10X | PBMC | Year 2 | 5' seq | no | yes |
| 4 | 1307_003_YR2_P | Wu_vdj_multi_MGI2498_10X | PBMC | Year 2 | 5' seq | yes | yes |
| 5 | 1307_004_YR2_P | Wu_vdj_multi_MGI2592_10X | PBMC | Year 2 | 5' seq | yes | yes |
| 6 | 1307_005_BSE_C | Wu_MGI0110_1_10X | CSF | Baseline | 5' seq | yes | yes |
| 7 | 1307_005_BSE_P | Wu_MGI0110_1_10X | PBMC | Baseline | 5' seq | yes | yes |
| 8 | 1307_005_YR1_C | Wu_MGI0818_10X | CSF | Year 1 | 5' seq | yes | yes |
| 9 | 1307_005_YR1_P | Wu_MGI0818_10X | PBMC | Year 1 | 5' seq | yes | yes |
| 10 | 1307_006_BSE_C | Wu_MGI0208_1_10X | CSF | Baseline | 5' seq | yes | yes |
| 11 | 1307_006_BSE_P | Wu_MGI0208_1_10X | PBMC | Baseline | 5' seq | yes | yes |
| 12 | 1307_006_YR1_C | Wu_vdj_multi_MGI2439_10X | CSF | Year 1 | 5' seq | yes | yes |
| 13 | 1307_006_YR1_P | Wu_vdj_multi_MGI2439_10X | PBMC | Year 1 | 5' seq | yes | yes |
| 14 | 1307_006_YR2_P | Wu_vdj_multi_MGI3572_10X | PBMC | Year 2 | 5' seq | yes | yes |
| 15 | 1307_007_W10_P | Wu_MGI0470_1_10X | PBMC | Week 10 | 5' seq | yes | yes |
| 16 | 1307_007_YR2_P | Wu_vdj_multi_MGI3167_10X | PBMC | Year 2 | 5' seq | no | yes |
| 17 | 1307_009_BSE_C | Wu_vdj_multi_MGI0964_10X | CSF | Baseline | 5' seq | no | yes |
| 18 | 1307_009_BSE_P | Wu_vdj_multi_MGI0964_10X | PBMC | Baseline | 5' seq | yes | yes |
| 19 | 1307_009_YR2_P | 1307_009_YR2_P | PBMC | Year 2 | 5' seq | yes | yes |
| 20 | 1307_010_BSE_P | Wu_MGI0884_10X | PBMC | Baseline | 5' seq | yes | yes |
| 21 | 1307_010_YR2_P | Wu_vdj_multi_MGI4051_10X | PBMC | Year 2 | 5' seq | yes | yes |
| 22 | 1307_011_BSE_C | Wu_vdj_multi_MGI1108_10X | CSF | Baseline | 5' seq | yes | Yes |
| 23 | 1307_011_BSE_P | Wu_vdj_multi_MGI1108_10X | PBMC | Baseline | 5' seq | no | yes |
| 24 | 1307_011_W10_C | Wu_vdj_multi_MGI2135_10X | CSF | Week 10 | 5' seq | yes | yes |
| 25 | 1307_011_W10_P | Wu_vdj_multi_MGI2135_10X | PBMC | Week 10 | 5' seq | yes | yes |
| 26 | 1307_011_YR2_P | MGI4305_TWHR-KLee-1307_011_YR2_P | PBMC | Year 2 | 5' seq | yes | yes |
| 27 | 1307_012_BSE_P | Wu_vdj_multi_MGI2004_10X | PBMC | Baseline | 5' seq | yes | Yes |
| 28 | 1307_012_YR2 | MGI4362_1307_012_YR2 | PBMC | Year 2 | 5' seq | yes | yes |
| 29 | 1307_013_BSE_C | Wu_vdj_multi_MGI2831_10X | CSF | Baseline | 5' seq | yes | yes |
| 30 | 1307_013_BSE_P | Wu_vdj_multi_MGI2831_10X | PBMC | Baseline | 5' seq | yes | yes |
| 31 | 1307_013_W05_C | Wu_vdj_multi_MGI2900_10X | CSF | Week 5 | 5' seq | yes | yes |
| 32 | 1307_013_W05_P | Wu_vdj_multi_MGI2900_10X | PBMC | Week 5 | 5' seq | yes | yes |
| 33 | 1307_0015_BSE_C | Wu_vdj_multi_MGI3541_10X | CSF | Baseline | 5' seq | yes | yes |
| 34 | 1307_0015_BSE_P | Wu_vdj_multi_MGI3541_10X | PBMC | Baseline | 5' seq | yes | yes |

## Supplemental Methods

### Sample processing

CSF samples were immediately processed upon acquisition. CSF samples were centrifuged at 400g for 10 minutes at 4°C and the cell pellets were resuspended in 400μL of CSF, and cell viability and counts were assessed using automated cell counters with viability dye. Heparinized blood was collected from patients, and peripheral blood mononuclear cells (PBMCs) isolated on ficoll density gradient. The CSF cell and PBMCs were resuspended at a concentration of 700-1000 cells/μL in PBS and 0.04% BSA and used to analyze the transcriptomic profile using single-cell RNA sequencing.

### Single-cell RNA-seq analysis

scRNA-seq reads were aligned to the GRCh38 reference genome and quantified using Cell Ranger count v.6.1.1. Filtered gene-barcode matrices that contained only barcodes with unique molecular identifier (UMI) counts that passed the threshold for cell detection were used for further analysis. QC analysis was performed using the Scanpy package v.1.9.1 in Python v.3.9.13 environment^1^. In each sample we filtered out cells: (1) with less than 200 genes, or greater than the 98% percentile; (2) with more than 10% mitochondrial transcripts; and (3) Red Blood Cell (RBC) clusters and platelets (defined as clusters that showed high expression of hemoglobin and platelet genes). Genes expressed in fewer than 10 cells were removed from the gene expression matrix. Cell doublets were determined using the Solo package^2^ v.1.2, as well as based on high expression levels of more than one canonical cell type-specific gene. Additionally, 1307_008_YR2_P was excluded from the analysis due to the low number of cells. Subsequently, all pre-processed samples were concatenated into a whole object. Next, data was normalized by a scale factor 10,000 and log transformed. Highly variable genes (HVGs) were selected using default parameters. The whole object was regressed out based on the total number of counts and the percentage of mitochondrial genes and subsequently scaled. The dataset was integrated by sample using the Harmony correction algorithm^3^ v.1.0. The shared nearest neighbor graph and UMAP were calculated using Harmony embeddings. For the neighbor graph, 40 principal components and 10 nearest neighbors were applied. Cell clusters were obtained by applying the Leiden algorithm in a wide range of resolutions and manually annotated with well-known markers to discriminate cell types.

### Cluster abundance analysis

To test the difference in cluster abundance (i.e. cell counts) between MS subjects at BSE and post-CladT treatment, we used the sccomp package^4^. The sccomp_estimate function was employed to calculate the differences in cell type composition using a linear model based on sum-constrained independent Beta-binomial distribution. The sccomp_remove_outliers function with default parameters was used to identify outliers and exclude them from the estimation. The sccomp_test function was employed to perform hypothesis testing. False-discovery rate (FDR) was calculated by sccomp package as described by Mangiola^4^ and FRD < 0.05 and FRD < 0.1 as well as log2FC > 0.5 was used to determine significance in cluster abundance in the PBMCs and CSF, respectively.

### Single-cell TCR analysis

Cell Ranger V(D)J Annotation pipeline v.6.1.1 was used for read alignment to the reference genome, GRCh38 (vdj_GRCh38_alts_ensembl-5.0.0) and to assemble TCRs. For downstream analysis, TCRs with only one pair of high-confidence and productive rearrangements for TCRα and TCRβ chains were selected. Clonotypes were defined based on identical CDR3 amino acid sequences from TCRα and TCRβ chains together. Gini coefficients in each sample were calculated using the Gini function from the DescTools package v.0.99.36. For clonal dynamics analysis the clonalCompare function from scRepertoire R package^5^ v.2.0.0 was applied.

### Prediction of viral-specific TCRs

We used a database of known TCR specificities established by published T cell specificity assays, VDJdb^6,7^ (https://vdjdb.cdr3.net) to determine the specificity of each TCR in our dataset. The VDJdb dataset was filtered for Human species clonotypes. Clonotypes were considered a match to a known viral-specific clonotype if the CDR3 sequence matched to the same viral epitope for either α- or β-chains of a TCR clonotype. No amino acid substitution was allowed between the sequenced and database TCRs.

### Single-cell BCR analysis

Cell Ranger V(D)J Annotation pipeline v.6.1.1 was used for read alignment to the reference genome, GRCh38 (vdj_GRCh38_alts_ensembl) and to assemble BCRs. For downstream analysis, BCRs with only one pair of high-confidence and productive rearrangements for light and heavy chains were selected. Cell Ranger output was converted into the AIRR format using changeo-10x superscript from the Immcantation framework (docker immcantation/suite:4.3.0) which re-assigned V, D, and J genes using the IMGT reference database of human alleles and reconstructed heavy chain germline V and J sequences. A sequence similarity threshold of 0.15 was chosen for clonotype grouping using the Hamming distance method. Clonotypes in each sample were grouped with DefineClones.py function from the Change-O package^8^ v1.2.0 based on 85% CDR-H3 nucleotide sequence similarity and identical allele-free IGHV and IGHJ usage. Gini coefficients in each sample were calculated using the Gini function from the DescTools package v.0.99.36.

### Analysis of SHM and Inference of B cell lineage trees

The SHM rate was quantified relative to the germline sequence of each V gene using the observedMutations function in the ShazaM package^8^ v.1.2.0. B cell lineage tree topologies and branch lengths were estimated within each subject using the dnaml program distributed as part of PHYLIP v.3.697. Trees were visualized using the R package ggtree^9^ v.3.11.2.

